# Face masks release water vapour but where does it go? An early observational study

**DOI:** 10.1101/2020.08.09.20154435

**Authors:** T.J. Stubington, R.S. Sahota, N. Mottacki, M.N. Johnston, O. Judd

## Abstract

**Objectives:** The aim of this observational study was to demonstrate the behaviour and trajectory of exhaled material from an individual wearing an FFP3 mask. Valves allow material release, but we theorised that valve design may direct material downwards towards patient and surrounding environment.

Limiting transmission of diseases with aerosolised spread is a current and serious concern within healthcare worldwide. Filtering face piece masks (FFP) are an essential piece of protective equipment when treating patients with ongoing infection. However, valved masks in other settings such as elective theatre and by the general public may have unforeseen negative effects.

**Design:** A heating coil-based vaporiser was used to produce visible water vapour. A healthy test subject was filmed wearing a variety of different masks and exhaling the water vapour.

**Results:** Flexible pleated and solid-shell FFP masks direct exhaled material downwards in plumes exceeding 25 cm. Duckbill-shaped masks appear to direct exhaled vapour laterally, with a smaller plume. The effect is influenced by mask design and type of valve. Fluid repellent surgical masks reduce material directed downwards, and when used in conjunction with an FFP3 mask, appear to reduce the size and density of the exhaled vapour plume. The use of a visor was ineffective in reducing plume expulsion.

**Interpretation:** A properly fit-tested FFP3-rated protective mask may only moderately limit expulsion of aerosolised particles from asymptomatic healthcare workers to patients, particularly in cases where procedures are being performed in close proximity to patients or in cases where mucosal surfaces are exposed. Further research in this area is needed.

## Introduction

Limiting transmission of diseases with aerosolised spread is a current and serious concern within healthcare worldwide. Recent developments – foremost among which is the emergence of the novel coronavirus SARS-CoV-2 – have led to the adoption of multiple strategies to limit the spread of infection in healthcare environments. These concerns, however, have been primarily focused on limiting transmission from patients to healthcare professionals. Three transmission routes of respiratory pathogens are described in the literature; (1) contact transmission (transmission through direct contact with a positive case or indirect contact via an object or surface), (2) droplet transmission (respiratory droplets from a positive case making contact with a mucosal surface), and (3) aerosol transmission (inhalation of small droplets that are suspended in the air as an aerosol)^3^. MERS, SARS, influenza viruses and a number of other organisms have been studied but there remains a dearth of high-quality evidence to clearly demonstrate the relative significance of each method of transmission^3,4^.

It has proven difficult to accurately quantify the infection vectors of virulent respiratory pathogens. Most responses to SARS-CoV-2 have included social restrictions intended to combat all 3 potential methods of transmission^5,6^. Self-isolation of vulnerable individuals, reductions in movements outside the home and a policy of maintaining distance between individuals reduces the opportunities for direct spread by touch. Rigorous cleaning of surfaces and hand washing reduces indirect spread. The wearing of water-resistant face masks limits the expulsion of respiratory droplets and hence droplet spread. Filtering face piece masks limit spread by inhalation by protecting those in direct contact with infected patients from aerosolised material which may contain the virus.

Previous pandemics such as SARS (caused by the coronavirus SARS-CoV-1) have demonstrated significant healthcare worker infection rates as a result of aerosol-generating procedures (AGPs)^7^. This trend is mirrored by COVID19 (caused by SARS-CoV-2), with rising numbers of healthcare worker deaths worldwide^8,9^. The National Health Service in the UK has taken extensive measures to prevent healthcare workers from contracting SARS-CoV-2 in the work setting.^8^ A key component of these measures is the provision of FFP masks. These protect the wearer from the external environment but particularly in the case of valved masks they do not protect the environment from the user’s exhaled material^10^. We present the results of an observational study which provides a visual representation of the material ejected from a user wearing a variety of masks, as well as the effect of valves in redirecting this material. We also provide a number of possible interventions to limit the negative impact of valved masks for those individuals unable to wear an alternative unvalved FFP3 mask.

## Methods

Four different FFP3 masks and a standard fluid-resistant surgical mask were tested. Each mask was produced by a different manufacturer. In the study they are described by a unique identifying letter for the duration of the paper. Table 1 shows the characteristics of each mask.

**Table 1:** This table describes the type of mask and relevant specific features and associates each mask with a unique identifying letter.

**Figure 1: Features and Descriptions of Each of the Tested Masks**
| **Mask ID** | **Protective Level** | **Design** | **Valve?** | **Nose clip?** | **Adjustable straps?** |
| --- | --- | --- | --- | --- | --- |
| **No mask** | **Nil** | **N/A** | **N/A** | **N/A** | **N/A** |
| **A** | Surgical mask | Fluid resistant surgical mask | No | No | No |
| **B** | FFP3 | Flexible pleated mask | Yes | Yes | No |
| **C** | FFP3 | Solid shell mask | Yes | Yes | Yes |
| **D** | FFP3 | Duck bill mask | Yes | Yes | No |
| **E** | FFP3 | Flexible pleated mask | No | Yes | No |

A healthy clean-shaven male test subject in his 20s (non-smoker, no known respiratory illnesses, no current medications, average peak expiratory flow of 720 l/min) was used to demonstrate the effects of each mask. The test subject had been appropriately fit tested to all of the masks. Fit testing was previously performed and complied with local hospital procedure. Prior to testing each of the masks were applied to the test subject’s face, adjusted and the fit was confirmed by sharp exhalation through the nose observing for misting of eyewear. A heating coil-based handheld vaporiser (Smok® RPM40, battery capacity 1500mAh, power set to 40 watts) was used to produce visible water vapour during exhalation. The test subject then fitted each mask in turn, lifted the base to introduce the vaporiser, held their breath while the mask fit was readjusted and then exhaled gently and passively over approximately 5 seconds. Following initial testing of each mask potential additional measures to mitigate valve output were also performed using the same test procedure (table 2). Video recordings were taken using an iPhone 11 Pro using the on-board 12-megapixel camera system (Apple Inc., California) against a plain blue background in a sheltered outdoor space with no exposure to wind. All recordings were taken on the same day in the same location, during sunshine and with no clouds overhead, to ensure consistent lighting during testing.

**Table 2:**
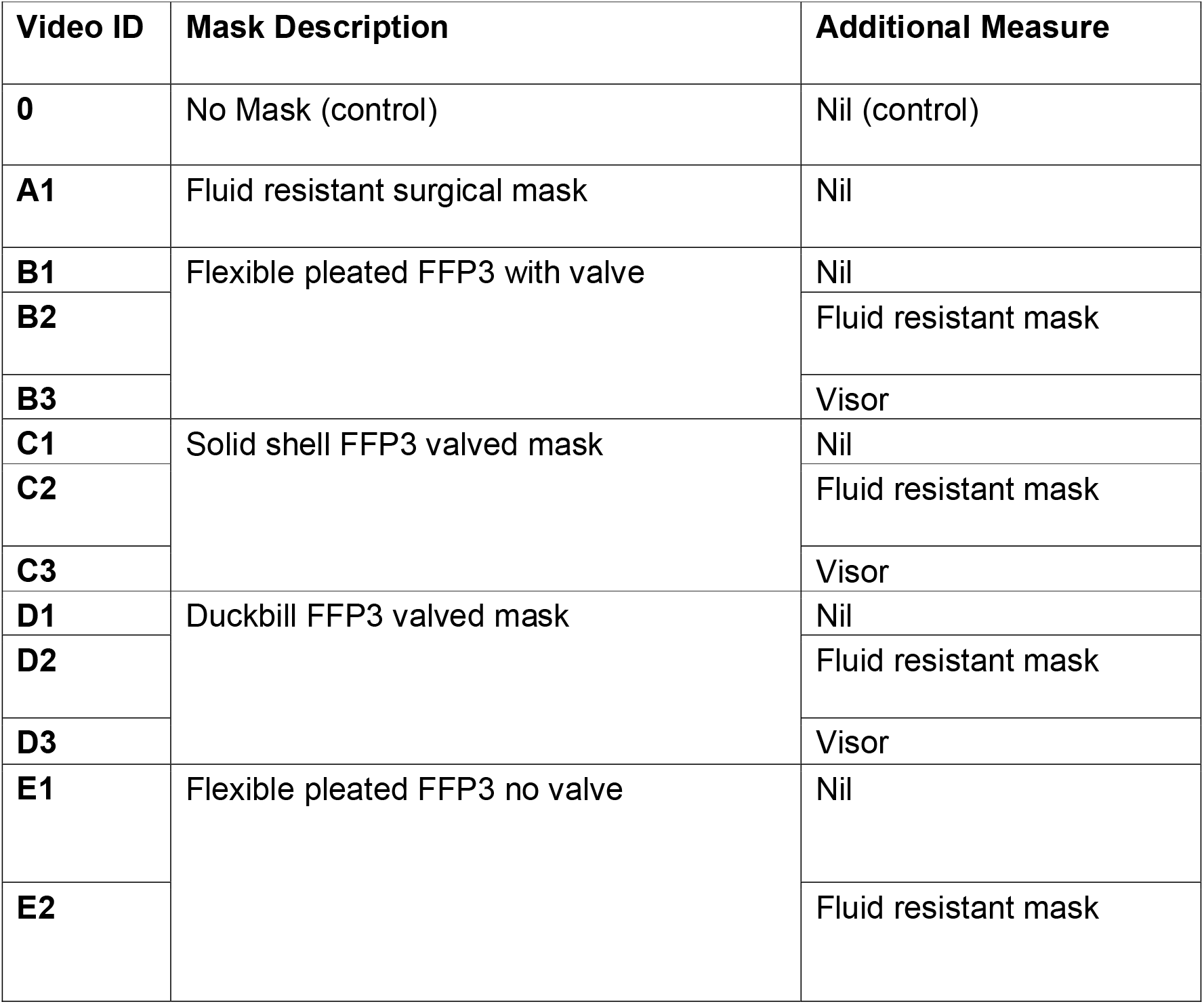
This table details the combinations of masks and additional measures tested and associates each combination with an alphanumerical combination. This alphanumerical code identifies the video recording of this test combination which is available in supplementary material.

## Results (Table 3)

### No mask (Table 3 row 0)

The image and video recording clearly demonstrate vapour flowing in an antero-inferior trajectory. The vapour travels in excess of 60 cm from the mouth of the subject (Table 3 Row 0).

**Table 3:**
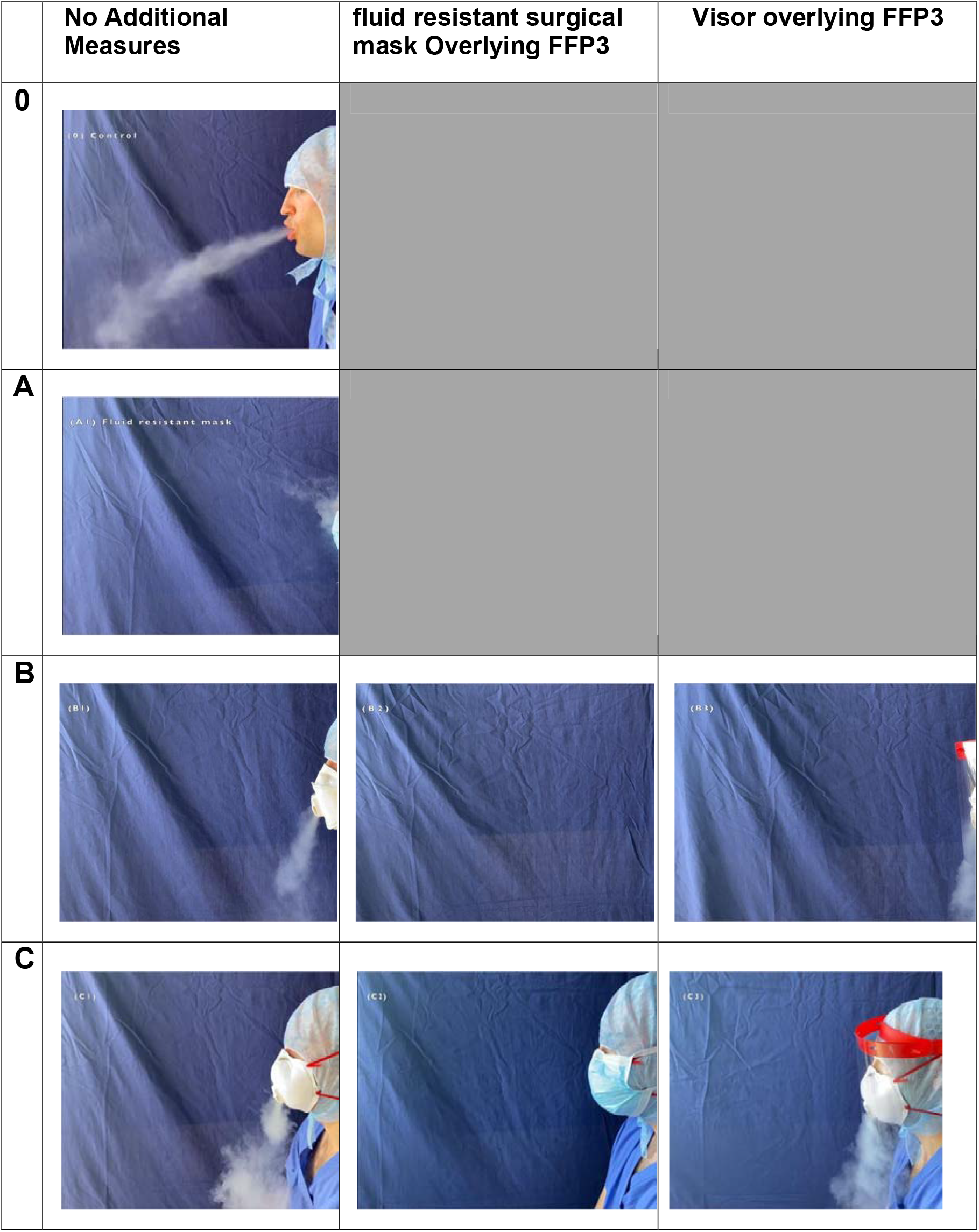
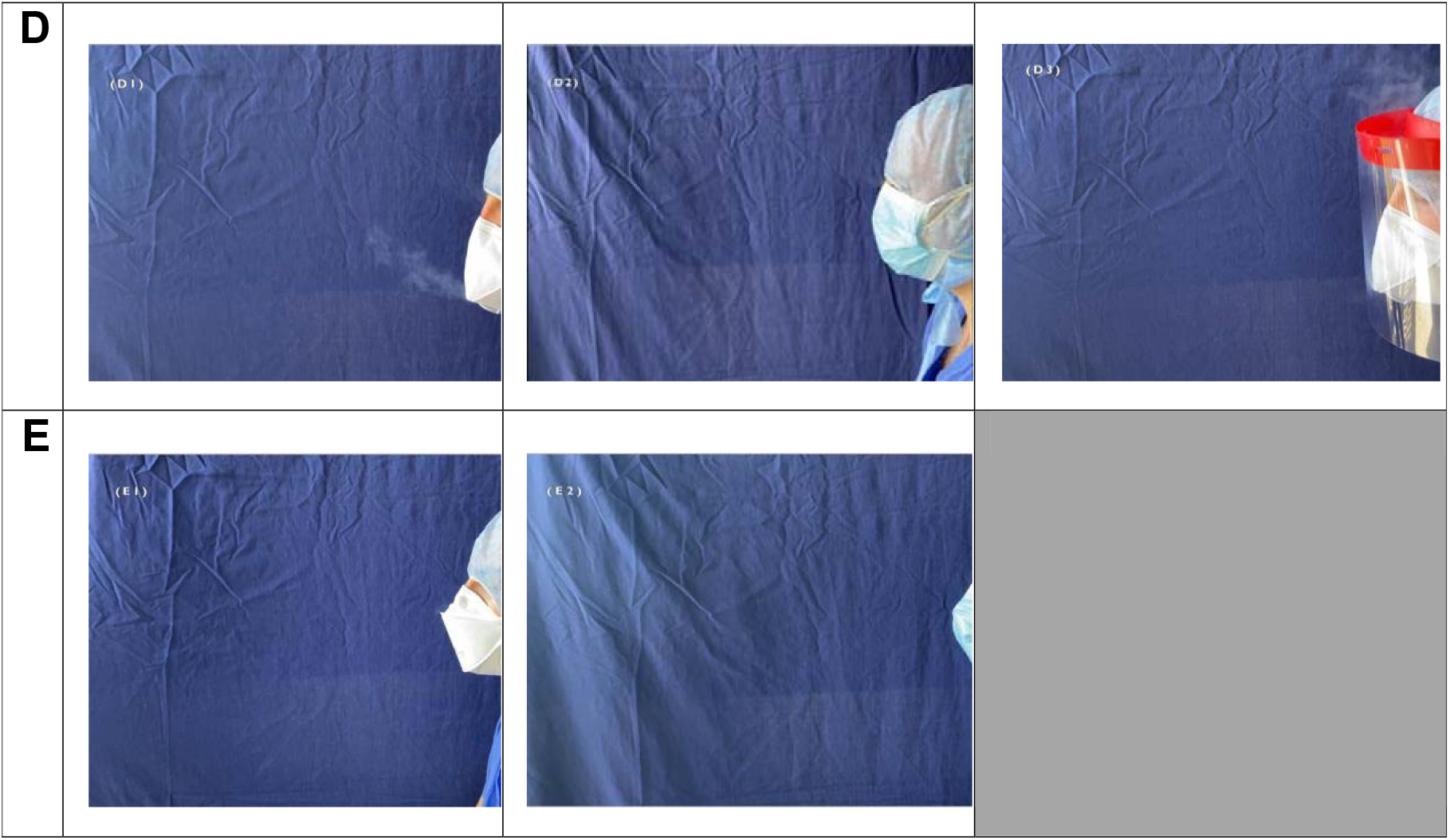
The stills taken from each experiment demonstrate the combinations of mask and additional measures and the effects that these combinations had on respiratory ejecta.

### Mask A (Table 3 row A)

The fluid resistant surgical mask was effective at directing exhaled vapour upwards and laterally while appearing to minimise inferior escape. There was no spread beyond 10cm from the subject which could be visualised as aerosol vapour (Table 3 Row A, A1).

### Mask B (Table 3 row B)

The valve in mask B allows exhaled vapour straight through the flapped valve system (B1). It directs the vapour downwards; maximum distance was measured at 25 cm from the valve outlet. Applying a fluid resistant surgical mask was effective in diminishing output (B2). The plume of vapour was reduced to only a few centimetres and redirected upwards. Applying a visor (B3) in conjunction with mask B allowed the vapour to concentrate. The thicker vapour was redirected downwards.

### Mask C (Table 3 row C)

The valve allows exhaled vapour through the flapped system, directing it downwards. The maximum spread of the plume was greater than 25 cm from the test subject. The addition of the fluid resistant surgical mask provides an effective additional barrier (C2), leading to minimal escape.

### Mask D (Table 3 row D)

Mask D directed the plume for aerosol forward and upwards with maximum extent of the visible plume around 12cm from the valve. When coupled with a fluid resistant mask (D2) minimal vapour was directed downwards and made the plume negligible. The use of a visor did not reduce the size and extent of the plume, instead predominantly directing the vapour upwards (D3).

### Mask E (Table 3 row E)

The valveless FFP3 mask limited the amount of vapour escaping (E1). A small amount escaped laterally, but was so insignificant as to make it difficult to quantify its size from the recordings. There was very little projected downwards. When a fluid resistant surgical mask was applied over mask E there was an additive effect with even less vapour ejecting and projecting downwards (E2). Since there appeared to be an insignificant plume, particularly upon adding a surgical mask, we did not proceed with testing the addition of a visor.

## Discussion

Our findings show that there is continued expulsion of aerosolised material despite the wearing of valved FFP3-rated masks. In the cases where our test subject wore flexible pleated or solid-shell masks, we observed the continued ejection of aerosol plumes downwards (figure 1). Applying a surgical mask on top of the FFP3 mask significantly reduces the size of the plume, and redirects it laterally in all cases (figure 2). The application of a visor appears to cause added deleterious effects to the direction and thickness of vapour. Though our data does not allow exact quantification of the expelled material, the vapour was projected downwards in excess of 25 cm from the valve in both cases. In the case of the solid-shell mask, there appears to be a greater amount of vapour being ejected; we hypothesise that this may be due to the rigid nature of the mask funnelling the exhalation. The addition of a visor had potentially negative effects causing increased downwards projection of the plume. We were curious if the application of tape on the exhaust of the solid-shell mask would impact the escape of vapour (figure 3). However, despite repeated attempts we found this was not an effective technique to seal the valve. There was significant variation between different tape applications, but none prevented vapour escape.

**Figure 1:**
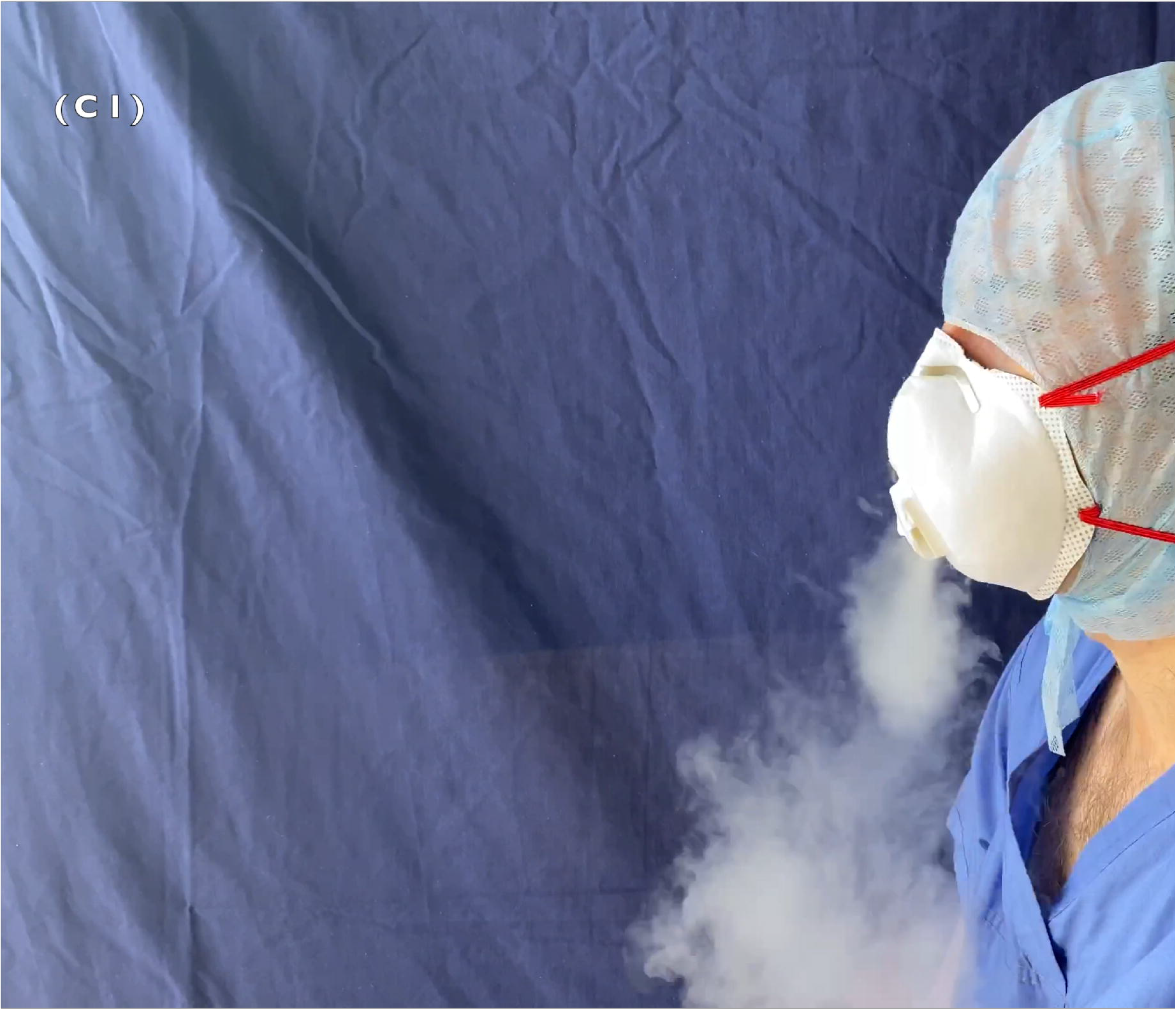
Valved mask with no additional measures in place (test C1)

**Figure 2:**
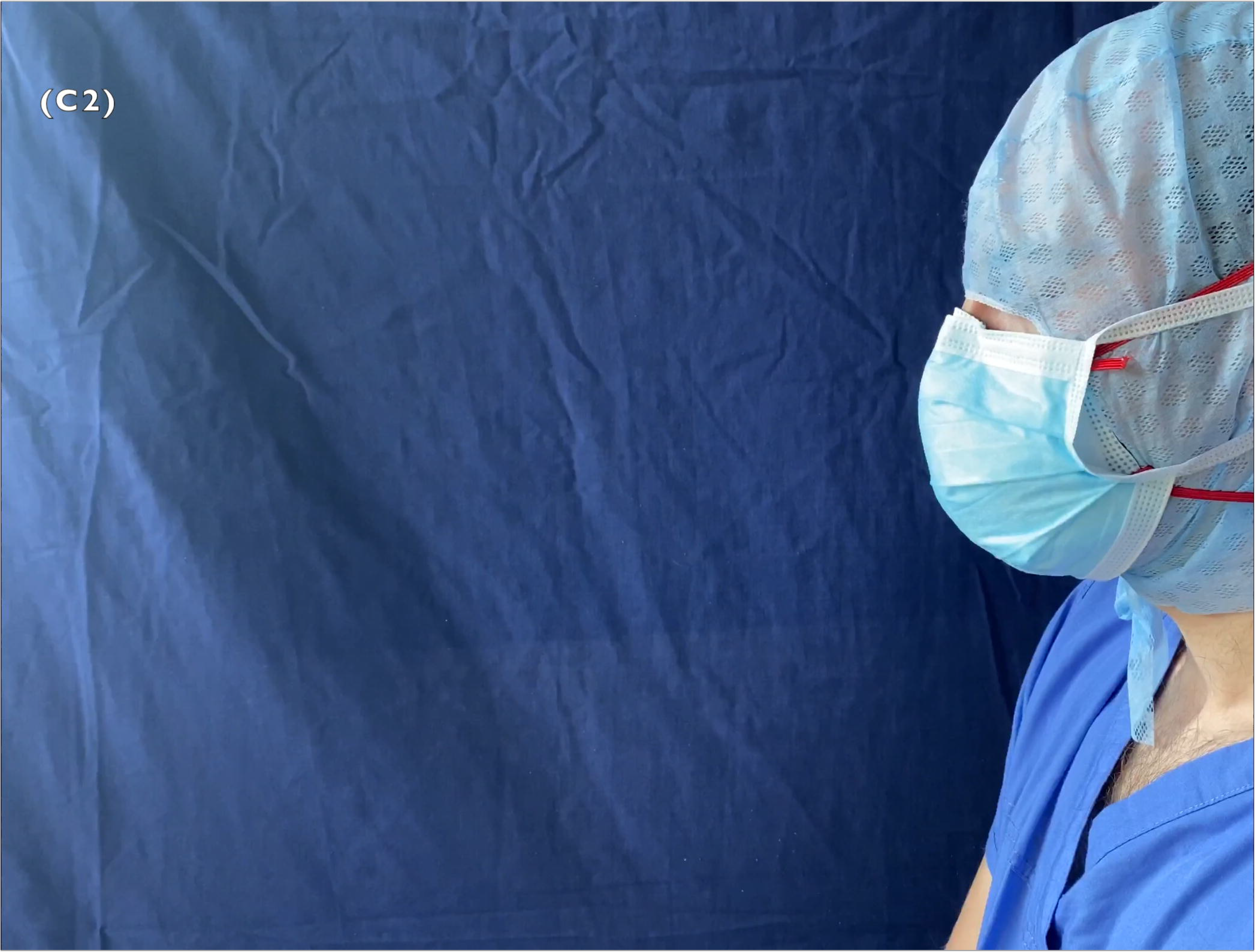
Valved mask with fluid resistant surgical mask placed over the top (test C2)

**Figure 3:**
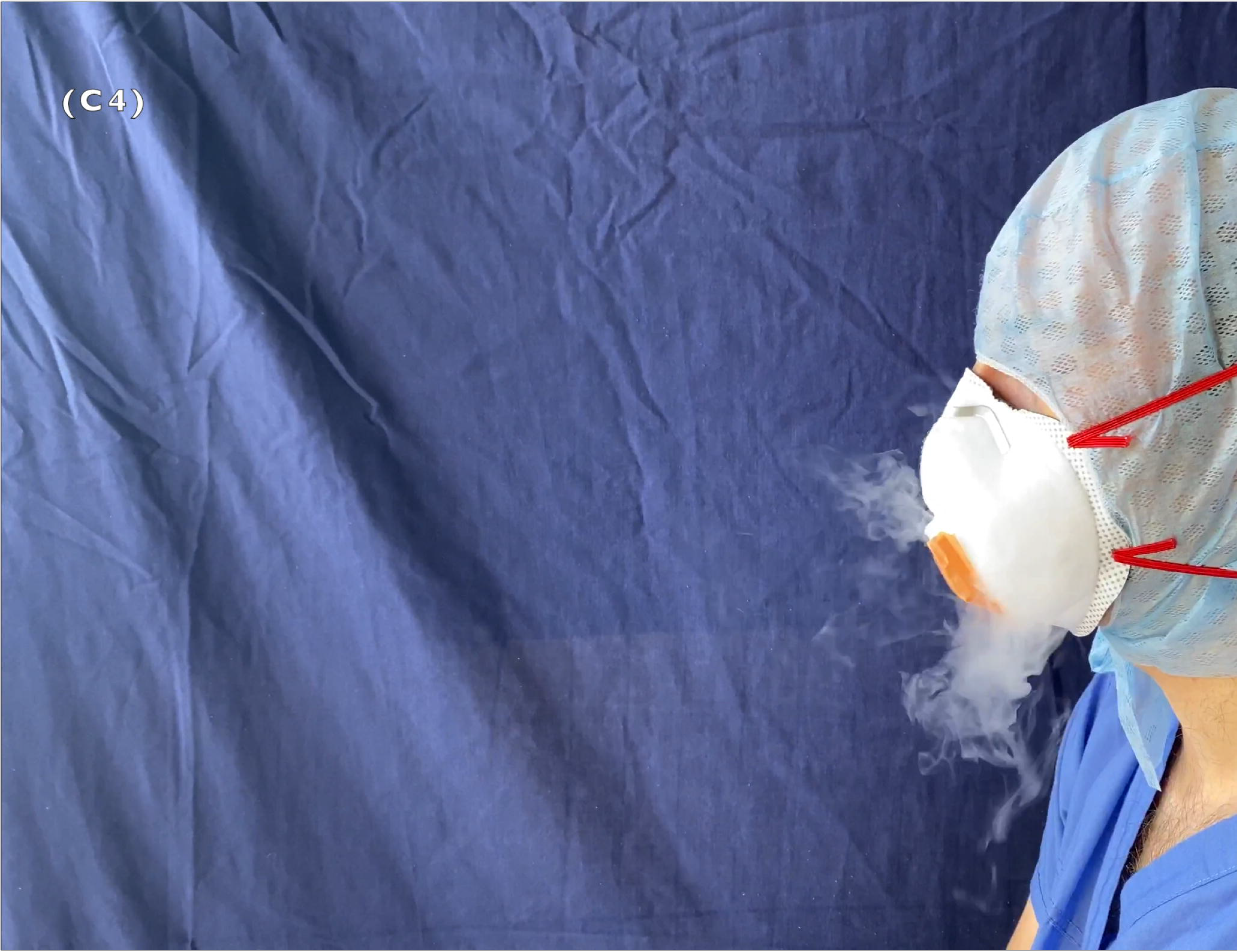
Tape applied over the valve of a valved mask (test C4). This proved ineffective as a solution to prevent exhaled material escaping.

Exhalation through the duckbill mask seemed to produce the smallest plume of the valved masks comparatively. These findings are not conclusive, but we hypothesise that the duckbilled shape of the mask diffuses aerosolised vapours prior to ejection.

This initial observational study is intended to provide a visual representation of a potential area of concern. Having observed the expulsion of droplets when using the recommended FFP3 masks, we have attempted to produce results to quantify the possibility of respiratory ejecta from healthcare workers using the currently recommended safety equipment. Further, this is highly relevant in the ongoing COVID-19 pandemic, with disease spread through aerosols having been demonstrated previously in other viruses^11-14^ and as yet unknown in SARS COV2^16^. Much of the response to the highly virulent SARS-CoV-2 virus has focused on limiting person to person spread^16^, though we have found no studies in the literature that assess the continued potential for aerosolised spread of the disease from healthcare workers to patients despite the use of protective equipment.

Viable respiratory viruses have been detected in droplets as small as 0.3μm and the size of droplets containing viable viruses seems to vary depending on the individual virus ^4,11-14^ The particles produced by the vaporiser are reported to be in the range of 10nm to 5μm^17,18^. Based on this size range and previously published data on other respiratory viruses it seems reasonable to conclude that droplets containing viable SARS-CoV2 could be released from these flutter valves into the surrounding environment.

Many countries around the world have made the wearing of face coverings mandatory in certain public places and within hospitals^19^. This has led the World Health Organisation supporting the use of non-medical face coverings in certain public settings despite a relative paucity of high quality evidence^16^. As urgent and elective activity is being reintroduced, most UK hospitals have strict protocols in place to protect patients and vulnerable healthcare workers^20-21^. It has been reported that asymptomatic or pre-symptomatic individuals can test positive for the virus and there have been reports of potential asymptomatic transmission^2,22-25^. It hospitals these masks are only recommended during aerosol generating procedures^16^. There are circumstances in which these findings, in conjunction with our results could be significant, such as if an asymptomatically positive healthcare worker wearing a valved mask is working in close proximity to a patient^22^. Although this is likely an uncommon occurrence, it has potentially catastrophic consequences for vulnerable patients, particularly those who are immunocompromised. Consideration should be given to the fact that the wearing of valved FFP3 masks to protect healthcare workers who are in contact with patients whose SARS-CoV-2 status is uncertain, represents a potential for continued spread of infection in the absence of more comprehensive precautions.

We are aware of the limitations of these results due to the absence of precision measurements, singular tests without repetition and variability in tidal volumes and force of exhalation. To an extent, these will have been minimised by using the same test subject and through attempting to standardise the evaluation protocol. It is difficult to draw definitive conclusions beyond comparing the vapour plume between the tests. In doing so, the images starkly demonstrate that in normal conditions, even in the absence of forced exhalation, the flutter valve permits exhaled material to enter the surrounding environment. Previous studies have demonstrated that fluid-repellent surgical masks are effective at directing exhaled material laterally and away from the patient^15,26^. Based on our results some FFP3 masks may instead be redirecting the exhaled material towards the patient. This may represent an increased risk of infection in cases where mucosal surfaces are exposed, e.g. during surgery or intubation. Further studies are mandatory to confirm and expand on our findings.

## Conclusion

Wearing FFP3-rated protective masks does not appear to eradicate the exhalation of aerosolised vapours. These findings may be relevant – in conjunction with modified protection procedures – to limit the expulsion of aerosolised particles from asymptomatic healthcare workers to patients, particularly in cases where procedures are being performed in close proximity to patients or in cases where mucosal surfaces are exposed. In our testing, duckbill-shaped masks appear to release the smallest amount of aerosolised particles; the addition of a surgical mask to the FFP3 masks appears to mitigate the amount of the plume, and also redirects it from the patient. The use of visors conveyed no such benefits. Our initial findings need to be corroborated by further research.

## Data Availability

The data from this paper in the form of video files is available from the corresponding author where it has not been included with the submission itself.

---

Supplementary video 1: Control, subject exhaling with no mask (test 0 as referenced in table 2)

Supplementary video 2: Water resistant surgical mask only (test A1 as referenced in table 2)

Supplementary video 3: Valved mask with no additional measures in place (test C1 as referenced in table 2)

Supplementary video 4: Valved mask with fluid resistant surgical mask over the top (test C2 as referenced in table 2)

Supplementary video 5: Valved mask with Visor (test C3 as referenced in table 2)

## Author Contributions

All credited authors were involved in the recording and acquisition of images

T Stubington wrote the majority of the article formatted the text for submission and prepared the non-photographic graphics for the paper

R Sahota arranged testing equipment, conducted an initial literature review and provided stylistic and content advice

N Mottacki wrote part of manuscript, providing stylistic and content advice

M Johnston sourced materials and constructed testing environment and provided review of manuscript for content and style

O Judd highlighted initial concerns, devised project, wrote initial introduction and reviewed manuscript for content and style as well as writing initial draft of conclusions.

## Role of Medical Writer

No medical writer was used at any point in the preparation of this manuscript all text was written by members of the research team.

## Patient and Other consent

No patients or patient information was involved in this study. The individual in the video footage and still images is the first author Mr T Stubington who has fully consented to the use of their image and the dissemination of recordings and images for all purposes related to the publication of this article. This study was approved by the local ethics committee at the Royal Derby Hospital.

## Supplementary information

Supplementary information is available for this paper in an online format. This comprises of the video recordings referenced in the text which can be viewed online.

## Acknowledgements

All of the credited authors are members of D.R.A.G.O.N the Derbyshire Rapid Aerosol Generating Observational Network. A research group set up by clinicians and researchers to explore aerosolisation using high speed video photography. This organisation is voluntary and no authors received any financial incentive to be a member nor has the D.R.A.G.O.N Organisation received any funding to date.

